# Factors associated with adherence to self-isolation and lockdown measures in the UK; a cross-sectional survey

**DOI:** 10.1101/2020.06.01.20119040

**Authors:** Louise E. Smith, Richard Amlôt, Helen Lambert, Isabel Oliver, Charlotte Robin, Lucy Yardley, G James Rubin

## Abstract

**Objectives:** To investigate factors associated with adherence to self-isolation and lockdown measures due to COVID-19 in the UK.

**Design:** Online cross-sectional survey.

**Setting:** Data were collected between 6^th^ and 7^th^ May 2020.

**Participants:** 2240 participants living in the UK aged 18 years or over. Participants were recruited from YouGov’s online research panel.

**Main outcome measures:** Having gone out in the last 24 hours in those who reported symptoms of COVID-19 in their household. Having gone out shopping for items other than groceries, toiletries or medicines (non-essentials), and total number of outings, in the last week in those who reported no symptoms of COVID-19 in their household.

**Results:** 217 people (9.7%) reported that they or someone in their household had symptoms of COVID-19 (cough or high temperature / fever) in the last seven days. Of these people, 75.1% had left the home in the last 24 hours (defined as non-adherent). Factors associated with non-adherence were being male, less worried about COVID-19, and perceiving a smaller risk of catching COVID-19. Adherence was associated with having received help from someone outside your household. Results should be taken with caution as there was no evidence for associations when controlling for multiple analyses. Of people reporting no symptoms in the household, 24.5% had gone out shopping for non-essentials in the last week (defined as non-adherent). Factors associated with non-adherence and with a higher total number of outings in the last week included decreased perceived effectiveness of Government “lockdown” measures, decreased perceived severity of COVID-19, and decreased estimates of how many other people were following lockdown rules. Having received help was associated with better adherence.

**Conclusions:** Adherence to self-isolation is poor. As we move into a new phase of contact tracing and self-isolation, it is essential that adherence is improved. Communications should aim to increase knowledge about actions to take when symptomatic or if you have been in contact with a possible COVID-19 case. They should also emphasise the risk of catching and spreading COVID-19 when out and about and the effectiveness of preventative measures. Using volunteer networks effectively to support people in isolation may promote adherence.

**WHAT IS ALREADY KNOWN ON THIS TOPIC:**

- The UK Government introduced “lockdown” measures, including physical or ‘social’ distancing, on 23^rd^ March 2020 due to COVID-19.
- Government guidance states that people with symptoms of COVID-19 should not leave their home, also known as self-isolation.
- There is no research investigating adherence to self-isolation and lockdown measures, or factors associated with self-isolation or lockdown measures in the UK.

**WHAT THIS STUDY ADDS:**

- Approximately 10% of participants indicated that they had had symptoms of potential COVID-19 (cough and high temperature / fever) in the last week. Of these participants, 75% had left their home in the last 24 hours.
- Factors associated with non-adherence to self-isolation measures included being male, less worried about COVID-19, and perceiving a smaller risk of catching COVID-19. However, these results should be taken with caution as there was no longer evidence for associations when correcting for multiple analyses.
- 25% of people who reported no symptoms in their household reported having gone out shopping for items other than groceries, toiletries or medicines in the last week; this was not allowed by Government guidelines in place at the time of data collection.
- Factors associated with non-adherence to lockdown measures, and increased number of outings in the last week, included decreased perceived effectiveness of Government “lockdown” measures, decreased perceived severity of COVID-19, and decreased estimates of how many other people were following lockdown rules.

## INTRODUCTION

On 23^rd^ March 2020, the UK Government introduced “lockdown” measures to slow the spread of COVID-19.(1, 2) These required people to stay at home except for: shopping for basic necessities (e.g. food or medicine), as infrequently as possible; exercising once a day; a medical need; or work purposes, but only those who could not work from home.(1) Guidance stated that while out and about, people should ensure that they were 2 metres apart from anyone outside their household. The requirements were different for households where anyone developed a continuous cough or fever. Those with symptoms themselves were required to stay at home, not leaving the home for any reason, for seven days from the start of their symptoms (“self-isolation”).(3) Those without symptoms, but who shared a home with someone who developed symptoms, were required not to leave the home for any reason for fourteen days from when the index case developed symptoms. Lockdown measures were extended on 16^th^ April (4) and eased slightly on 11^th^ May 2020 (5, 6), however no changes have been made to the requirements for people who developed potential symptoms of COVID-19.

Adherence to these measures is likely to be influenced by multiple factors. There is some evidence that people who think they have had COVID-19 are less likely to adhere to lockdown measures.(7) According to Protection Motivation Theory (8) uptake of a protective behaviour is influenced by your appraisal of a threat, including perceptions about its severity and your susceptibility to it, and your appraisal of the behaviour, including: perceptions about its efficacy, your ability to perform the behaviour, and the costs associated with the behaviour. A recent rapid review of factors affecting adherence to quarantine measures in previous public health crises found that knowledge of quarantine measures and perceived social norms were also associated with adherence to quarantine.(9) Conversely, fear of missing out, perceived social pressure, perceived legal consequences, running out of supplies (e.g. food or medicine) and financial pressures were associated with decreased adherence.

In this study, we investigated factors associated with adherence to self-isolation and lockdown measures in a demographically representative sample of the UK adult population. In households that reported symptoms of COVID-19, we investigated psychological, situational, personal and clinical factors associated with having left the home in the past 24 hours. In households where no symptoms of COVID-19 were reported, we investigated psychological, situational, personal and clinical factors associated with adherence to one of the UK lockdown rules (not going out to shop for items other than groceries, toiletries or medicines) and with total number of outings in the last seven days.

## METHOD

### Design

We commissioned the market research company YouGov to carry out this cross-sectional survey, between 6^th^ and 7^th^ May 2020.

### Participants

Participants (n = 2240) were recruited from YouGov’s online research panel (n = 800,000+ UK adults) and were eligible for the study if they were aged eighteen years or over and lived in the UK. Quota sampling was used, based on age, gender, social grade, highest level of education, and Government Office Region, to ensure that the sample was broadly representative of the UK general population. Online quota sampling is a standard approach used in opinion polling and can be a pragmatic approach when a large, demographically representative sample must be obtained in a very short time frame, particularly during a crisis.(10–13) While these surveys generally have low response rates, the risk of response bias is offset by the use of quotas to ensure demographic representativeness. The use of quotas also means that the ‘response rate’ is not a good indicator of response bias. Potential participants belonging to quotas that have already been filled are prevented from completing the survey. Of 2,623 people who began the survey, 2,314 completed it. Seventy-four participants were not included in the sample due to a lack of data for sociodemographic variables (meaning that they could not be assigned to a quota), suspiciously speedy completion of the survey (“speeding”) or providing identical answers to multiple consecutive questions (“straight-lining”). Participants were reimbursed in points (equivalent to approximately 50p) which could be redeemed as cash, gift vouchers or charitable donations.

### Study materials

Full survey materials are available in the supplementary materials.

#### Outcome measures

We asked participants to state how many times they had left their home “in the past 24 hours” and “in the past seven days”: to go to the shops for groceries, toiletries or medicine; to go to the shops for other items; for exercise; for a medical purpose excluding going to the shops/pharmacy for medicine; to go to work; to help someone else; and to meet friends or family who they did not live with.

#### Psychological and situational factors

We asked participants if they had experienced symptoms “in the past seven days” from a list of thirteen symptoms including cough and high temperature / fever (for full list see supplementary materials). We asked participants who lived with someone else if “someone else in [their] household” had experienced symptoms “in the past fourteen days” from the same list of thirteen symptoms.

We asked participants whether they thought they had “had, or currently have, coronavirus”. Possible answers were “I have definitely had it or definitely have it now”, “I have probably had it or probably have it now”, “I have probably not had it and probably don’t have it now”, and “I have definitely not had it and definitely don’t have it now”.

We asked if participants were currently self-isolating. Response options were “not self-isolating”, “self-isolating for seven days”, “self-isolating for fourteen days”, and “self-isolating for at least 12 weeks”. Twelve weeks was offered as an option because of UK guidance asking some extremely clinically vulnerable people to remain at home and ‘shield’ themselves for this length of time. People in this category were alerted to do this via a letter sent from the National Health Service.

We asked participants a series of true / false statements about UK Government guidance. Statements referred to households where no-one had symptoms or was extremely clinically vulnerable and where someone had symptoms of COVID-19 and no-one was extremely clinically vulnerable (all statements were false under these conditions). See supplementary materials for full item list.

We asked participants how worried they were about COVID-19 on a five-point Likert-type scale from “not at all worried” to “extremely worried”.

To measure perceived social norms, we asked participants to estimate the percentage of people the same age as them who were fully following the UK Government’s recommendations to stay at home.

We asked participants whether they thought the current lockdown had made their physical health better or worse. Possible answers were “a lot better”, “a little better”, “no difference”, “a little worse”, and “a lot worse”.

We asked participants to rate their general health on a five-point Likert-type scale from “poor” to “excellent” using one item from the SF-36.(14)

We asked participants if they had helped someone, or received help from someone, outside their household in the past seven days (yes / no).

We asked participants to rate fourteen perception statements on a five-point Likert scale from “strongly disagree” to “strongly agree”. Statements included the perceived severity of COVID-19, perceived effectiveness of Government measures, perceived likelihood of catching and spreading COVID-19, perceived costs of following Government measures, fear of losing touch with friends and relatives, social pressure from friends and family to follow Government measures, perceived legal consequences of not following Government measures, and positive consequences of the lockdown (see supplementary materials for full item list).

#### Personal and clinical characteristics

We asked participants to report their age, gender, employment status, highest educational or professional qualification, and their marital status. We also asked participants whether there was a child in their household, whether they or someone else in their household received a letter from the National Health Service telling them they were extremely clinically vulnerable to COVID-19, and whether they lived alone. Participants were asked for their postcode to determine indices of multiple deprivation (IMD) and whether they lived in an urban or rural area. We also collected social grade.

We asked participants if their primary home had access to any outdoor space, and whether they were pet owners.

### Ethics

Ethical approval for this study was granted by the King’s College London Research Ethics Committee (reference: LRS-19/20-18687).

### Patient and public involvement

Due to the rapid nature of this research, the public was not involved in the development of the survey materials.

### Power

We calculated achieved power for the analyses (in households with and without symptoms) using post-hoc power calculations. Achieved power is presented underneath relevant analyses.

### Analysis

Recoding of variables

We created a binary variable indicating whether participants had been out for any reason in the last 24 hours. Among those who reported symptoms in their household in the last seven days, we defined those who reported having gone out in the last 24 hours as not adhering to self-isolation measures.

We calculated the total number of outings made in the last seven days by summing the number of times people reported going out (shopping for groceries, toiletries or medicine, shopping for other items, exercising, for a medical purpose, going to work, helping someone else, meeting friends or family). Some participants (n = 54, 2.4%) reported going out many times and we capped this at twenty times in the past seven days. Based on UK Government guidelines which were in force at the time of data collection (1) we recoded going to the shops for items other than groceries, toiletries or medicine, meeting friends or family that did not live in your household, and having had visitors to your home in the past seven days as not adhering to UK Government guidelines.

We recoded presence of household symptoms in the last seven days into a binary variable. Presence of symptoms was defined as a participant reporting that they experienced cough or a high temperature / fever in the last seven days, or if a member of their household had experienced cough or a high temperature / fever in the last fourteen days. We recoded answers of “don’t know” as missing (n = 13 own symptoms, n = 42 household member symptoms). Participants were able to skip questions about symptoms, resulting in some missing data (n = 36 own symptoms, n = 7 household member symptoms). Where participants indicated that they themselves had symptoms, but data for household member symptoms was missing, we recoded these data as symptoms being present in the household.

We recoded whether participants thought they had had, or currently had, COVID-19 into a binary variable, by grouping answers of “probably” and “definitely” had or have coronavirus, and “probably not” and “definitely not” had or have coronavirus.

We recoded whether participants were currently self-isolating into a binary variable (not self-isolating, self-isolating for any length of time).

We recoded knowledge of UK Government guidelines into two binary variables, one for if no-one in the household had symptoms, and one for if someone in the household had symptoms. Participants who identified all aspects of the guidance correctly were coded as being correct; those who answered any item incorrectly or did not know were coded as incorrect/unsure.

For all variables, unless stated otherwise, we coded answers of “don’t know” as missing data.

#### Analyses

We investigated whether the total number of outings differed by presence of symptoms in the household by using an independent samples t-test. We investigated whether the percentage of people reporting going out shopping for items other than groceries, toiletries or medicines (non-essentials), going out to meet up with friends or family and having visitors to their home differed by presence of symptoms in the household by using chi-squared tests.

We split the sample by presence of symptoms in the household. We tested factors associated with non-adherence to self-isolation measures in those who reported symptoms in the household. We ran a series of logistic regressions investigating univariable associations between personal and clinical factors, psychological and situational factors and having left the home in the past 24 hours. We ran a second set of logistic regressions controlling for personal and clinical characteristics (gender, age, having a child in the household, being extremely clinically vulnerable, employment status, highest educational or professional qualification, indices of multiple deprivation, social grade, living in a rural or urban area, living alone, marital status, and region).

We investigated factors associated with non-adherence to lockdown measures in those who reported no symptoms in the household. We ran a series of linear regressions investigating univariable associations between personal and clinical factors, psychological and situational factors and total number of outings reported in the past seven days (same variables as above). We ran a second set of linear regressions controlling for personal and clinical characteristics. For these analyses, we entered personal and clinical characteristics as the first block, and other independent variables as the second block. We used the total proportion of the variance explained by individual psychological and situational factors and personal and clinical characteristics, and statistical significance of individual regression coefficients to determine the psychological and situational factors most influential in number of outings in the past week. We ran a series of logistic regressions investigating univariable associations between personal and clinical factors, psychological and situational factors and going out shopping for items other than groceries, toiletries or medicines in the past seven days. We ran a second set of logistic regressions controlling for personal and clinical characteristics (same variables as above).

Weighting data by age, gender, social grade, highest level of education, and region altered prevalence of outcome behaviours only slightly (1.1% for having been out in the last 24 hours, and 0.4% for going out shopping for non-essentials; mean difference in number of outings in past week 0.15). We therefore used unweighted data in our analyses.

#### Sensitivity analyses

Due to the large number of analyses (n = 39) run on each outcome, we applied a Bonferroni correction to our results (*p*≤.001). Those meeting this criterion are marked by a double asterisk (**) in the tables.

## RESULTS

A minority of participants (9.7%, n = 217) reported that either they or a household member had a cough or a high temperature / fever in the last seven days. Participant characteristics are shown in Table 1. Male participants were more likely to report symptoms in their household. There were no other differences between groups.

**Table 1.**
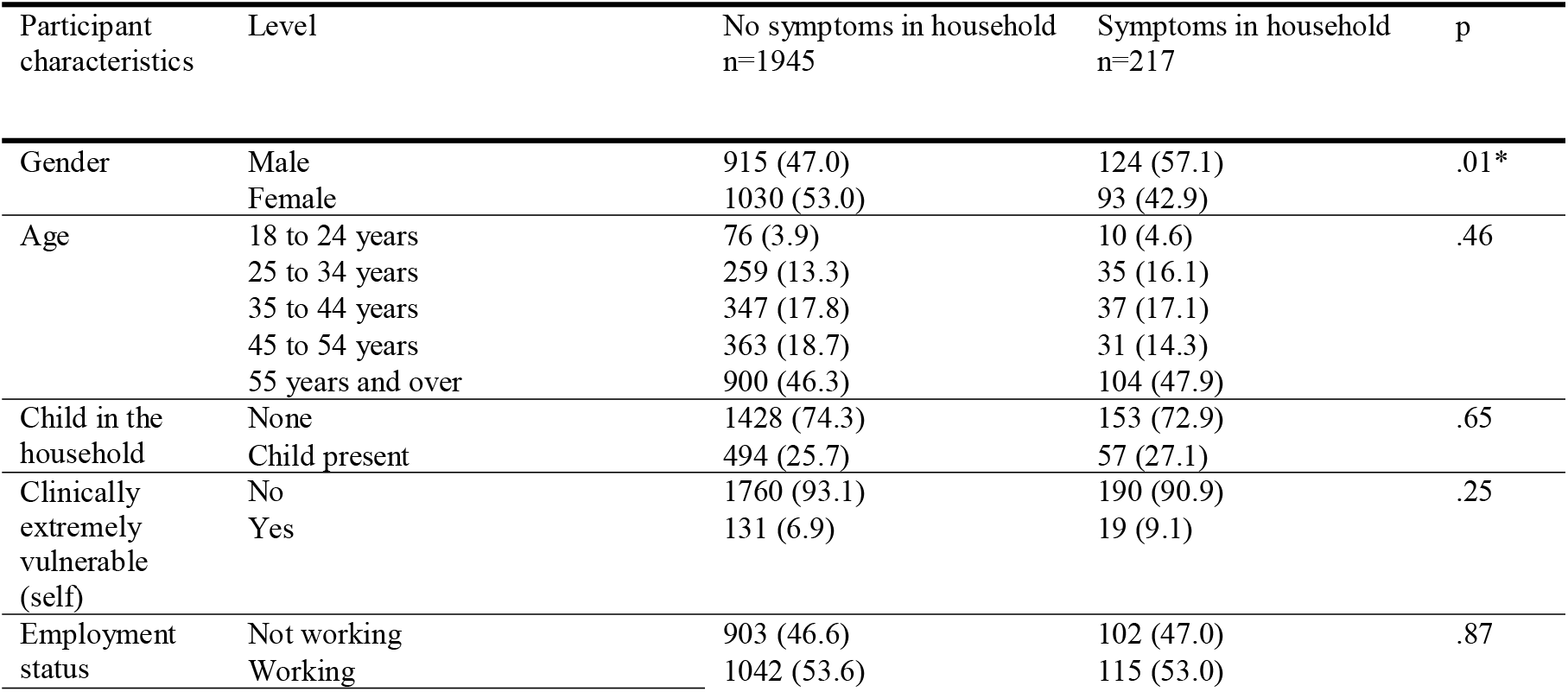
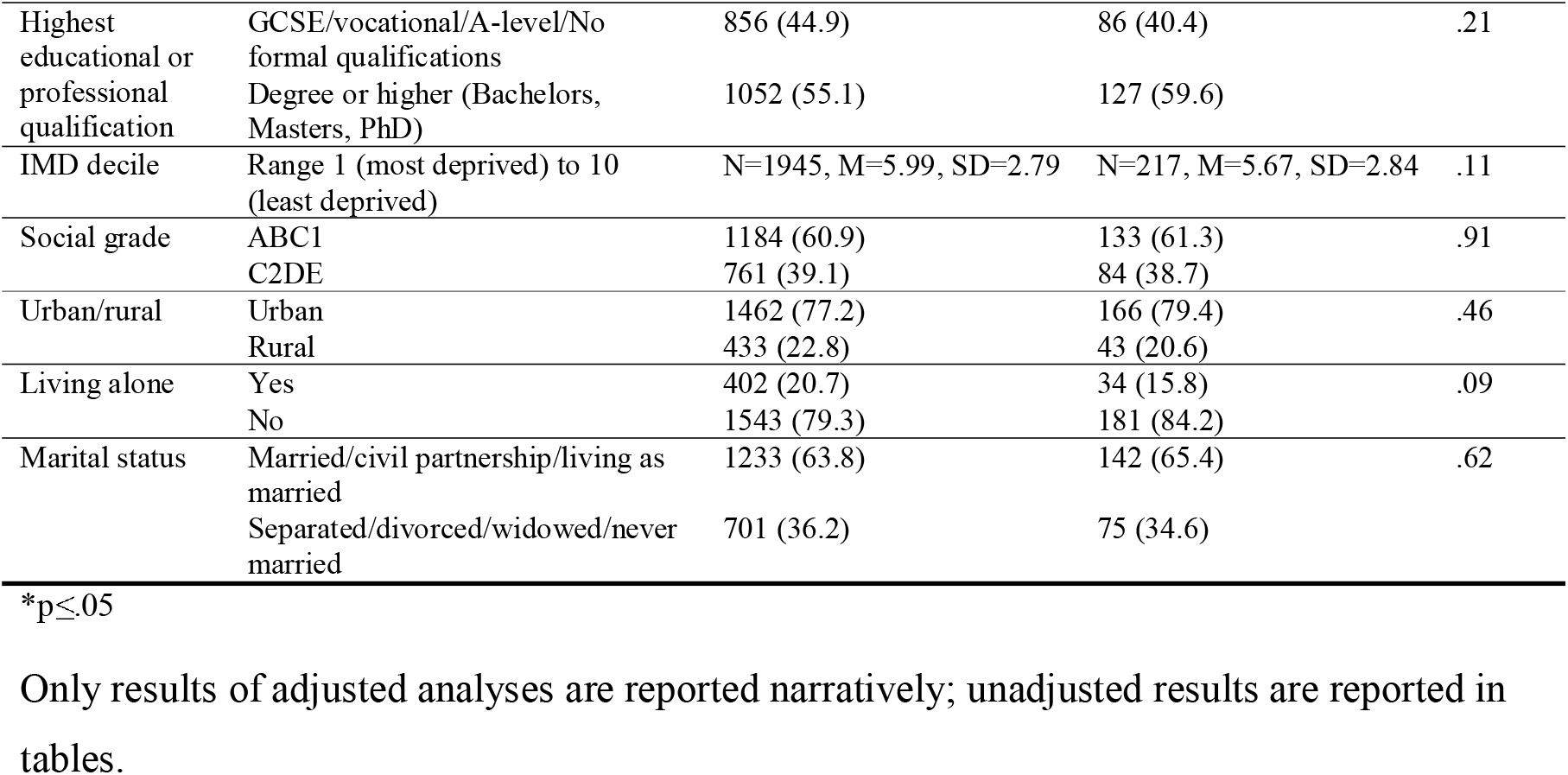
Participant personal and clinical characteristics, by report of symptoms in the household.

Only results of adjusted analyses are reported narratively; unadjusted results are reported in tables.

### Symptoms in household

Of participants who reported symptoms in their household (n = 217), 75.1% (n = 163, 95% CI [69.3 to 80.9]) reported leaving the home at least once in the past 24 hours. This finding has been reported elsewhere.(15)

When we compared participants with symptoms in the household versus those without symptoms we found no differences in the total number of outings made in the last week (*t*(2160) = 0.20, *p* = .84), nor the percentage of people reporting having gone out shopping for items other than groceries, toiletries or medicines (χ^2^(1, 2162) = 0.38, *p* = .54), having had a visitor to their home (χ^2^(1, 2076) = 0.40, *p* = .53), or having gone out to meet friends or family they did not live with (χ^2^(1, 2162) = 1.34, *p* = .25).

Of those who reported symptoms in the household, 34.1% (n = 74) reported self-isolating either for seven days (n = 19), fourteen days (n = 13), or at least 12 weeks (n = 42). However, of those that described themselves as self-isolating, 60.8% (n = 45) nonetheless reported having gone out in the last 24 hours.

In terms of personal and clinical characteristics, leaving the home in the last 24 hours was associated with being male (see Table 2).

**Table 2.** Associations between personal and clinical characteristics of participants who reported symptoms in their household in the last week and having left the home in the past seven days.

| Participant characteristics | Level | Did not go out in past 24 hours<br>n=54 | Went out in past 24 hours<br>n=163 | Odds ratio<br>(95% CI) | Adjusted odds ratio<br>(95% CI)<sup>†</sup> |
| --- | --- | --- | --- | --- | --- |
| Gender | Male | 23 (18.5) | 101 (81.5) | Reference | Reference |
|  | Female | 31 (33.3) | 62 (66.7) | 0.46 (0.24 to 0.85)* | 0.33 (0.14 to 0.77)* |
| Age | 18 to 24 years | 4 (40.0) | 6 (60.0) | Reference | Reference |
|  | 25 to 34 years | 8 (22.9) | 27 (77.1) | 2.25 (0.51 to 9.99) | 1.98 (0.26 to 15.38) |
|  | 35 to 44 years | 6 (16.2) | 31 (83.8) | 3.44 (0.74 to 16.03) | 2.05 (0.26 to 16.11) |
|  | 45 to 54 years | 10 (32.3) | 21 (67.7) | 1.40 (0.32 to 6.10) | 0.99 (0.14 to 6.82) |
|  | 55 years and over | 54 (24.9) | 163 (75.1) | 2 (0.52 to 7.64) | 2.09 (0.29 to 14.89) |
| Child in the household | None | 44 (28.8) | 109 (71.2) | Reference | Reference |
|  | Child present | 7 (12.3) | 50 (87.7) | 2.88 (1.21 to 6.85)* | 2.82 (0.88 to 9.01) |
| Clinically extremely vulnerable (self) | No | 44 (23.2) | 146 (76.8) | Reference | Reference |
|  | Yes | 8 (42.1) | 11 (57.9) | 0.41 (0.16 to 1.09) | 0.40 (0.10 to 1.55) |
| Employment status | Not working | 34 (33.3) | 68 (66.7) | Reference | Reference |
|  | Working | 20 (17.4) | 95 (82.6) | 2.37 (1.26 to 4.48)* | 2.62 (0.97 to 7.10) |
| Highest educational or professional qualification | GCSE/vocational/A-level/No formal qualifications | 22 (25.6) | 64 (74.4) | Reference | Reference |
|  | Degree or higher (Bachelors, Masters, PhD) | 29 (22.8) | 98 (77.2) | 1.16 (0.61 to 2.20) | 0.73 (0.29 to 1.82) |
| IMD decile | Range 1 (most deprived) to 10 (least deprived) | N=54,<br>M=5.81,<br>SD=2.69 | N=163,<br>M=5.63,<br>SD=2.89 | 0.98 (0.88 to 1.09) | 1.08 (0.91 to 1.27) |
| Social grade | ABC1 | 37 (27.8) | 96 (72.2) | Reference | Reference |
|  | C2DE | 17 (20.2) | 67 (79.8) | 1.52 (0.79 to 2.92) | 2.37 (0.89 to 6.32) |
| Urban/rural | Urban | 39 (23.5) | 127 (76.5) | Reference | Reference |
|  | Rural | 12 (27.9) | 31 (72.1) | 0.79 (0.37 to 1.69) | 0.63 (0.22 to 1.80) |
| Living alone | Yes | 11 (32.4) | 23 (67.6) | Reference | Reference |
|  | No | 41 (22.7) | 140 (77.3) | 1.63 (0.73 to 3.63) | 0.50 (0.12 to 2.09) |
| Marital status | Married/civil partnership/living as married | 27 (19.0) | 115 (81.0) | Reference | Reference |
|  | Separated/divorced/widowed/never married | 27 (36.0) | 48 (64.0) | 0.42 (0.22 to 0.78)* | 0.39 (0.11 to 1.33) |
| Clinically extremely vulnerable (household member)<sup>‡</sup> | No | 38 (23.6) | 123 (76.4) | Reference | Reference |
|  | Yes | 3 (21.4) | 11 (78.6) | 1.13 (0.30 to 4.27) | 3.79 (0.51 to 28.07) |
| Home includes access to outside space | No | 4 (36.4) | 7 (63.6) | Reference | Reference |
|  | Yes | 50 (24.3) | 156 (75.7) | 1.78 (0.50 to 6.34) | 0.40 (0.04 to 4.48) |
| Pet ownership | No | 31 (31.6) | 67 (68.4) | Reference | Reference |
|  | Yes | 23 (19.3) | 96 (80.7) | 1.93 (1.04 to 3.60)* | 1.66 (0.70 to 3.97) |

\* $p \leq .05$
<sup>†</sup> Adjusting for gender, age, having a child in the household, being extremely clinically vulnerable oneself, employment status, highest level of education or professional qualification, indices of multiple deprivation, social grade, living in a rural or urban area, living alone, marital status, and region.
<sup>‡</sup> Adjusted analyses for this variable did not control for living alone, as by definition all participants asked this question lived in a household with someone else.

Leaving the home in the last 24 hours was associated with: thinking that the current lockdown had made your mental health worse; having better self-reported general health; and feeling a greater sense of community with your neighbourhood due to COVID-19 (see Table 3). Not leaving the home in the last 24 hours was associated with: reporting that you were self-isolating; increased worry about COVID-19; having received help from someone outside your household in the last seven days because of COVID-19; and increased perceived likelihood of catching COVID-19.

**Table 3.** Associations between psychological and situational factors and having left the home in the past seven days in participants who reported symptoms in the household.

| Participant characteristics | Level | Did not go out in past 24 hours n=54 | Went out in past 24 hours n=163 | Odds ratio (95% CI) | Adjusted odds ratio (95% CI)† |
| --- | --- | --- | --- | --- | --- |
| Had, or currently have, COVID-19 | Think have not had COVID-19 and do not have it now | 27 (20.8) | 103 (79.2) | Reference | Reference |
|  | Think have had COVID-19 or have it now | 17 (37.0) | 29 (63.0) | 0.45 (0.21 to 0.93)* | 0.31 (0.08 to 1.11) |
| Self-isolating | Not self-isolating | 25 (17.5) | 118 (82.5) | Reference | Reference |
|  | Self-isolating | 29 (39.2) | 45 (60.8) | 0.33 (0.17 to 0.62)** | 0.24 (0.09 to 0.62)* |
| Understanding of Government measures if no-one in household was symptomatic | Incorrect / unsure | 34 (26.6) | 94 (73.4) | Reference | Reference |
|  | Correct | 20 (22.5) | 69 (77.5) | 1.25 (0.66 to 2.35) | 0.95 (0.40 to 2.22) |
| Understanding of Government measures if someone in household was symptomatic | Incorrect / unsure | 49 (24.1) | 154 (75.9) | Reference | Reference |
|  | Correct | 5 (35.7) | 9 (64.3) | 0.57 (0.18 to 1.79) | 1.36 (0.30 to 6.22) |
| Worry about COVID-19 | 5-point scale, 1=not at all worried to 5=extremely worried | N=54, M=3.70, SD=0.92 | N=163, M=3.44, SD=1.02 | 0.77 (0.56 to 1.05) | 0.61 (0.37 to 0.98)* |
| Perceived social norms | Percentage (range 0-100) | N=46, M=72.13, SD=20.53 | N=151, M=69.84, SD=17.39 | 0.99 (0.97 to 1.01) | 0.99 (0.97 to 1.02) |
| Perceptions about impact on mental health | 5-point scale, 1=a lot better to 5=a lot worse | N=54, M=3.37, SD=1.07 | N=160, M=3.57, SD=0.96 | 1.22 (0.90 to 1.67) | 1.64 (1.05 to 2.56)* |
| Perceptions about impact on physical health | 5-point scale, 1=a lot better to 5=a lot worse | N=54, M=3.54, SD=0.91 | N=162, M=3.38, SD=0.91 | 0.82 (0.58 to 1.16) | 0.78 (0.48 to 1.27) |
| Self-reported general health | 5-point scale, 1=poor to 5=excellent | N=54, M=2.33, SD=1.13 | N=161, M=2.75, SD=0.96 | 1.51 (1.10 to 2.06)* | 1.56 (1.00 to 2.41)* |
| Helped someone outside household | No | 44 (28.6) | 110 (71.4) | Reference | Reference |
|  | Yes | 9 (14.8) | 52 (85.2) | 2.31 (1.05 to 5.09)* | 2.53 (0.91 to 7.05) |
| Received help from someone outside household | No | 37 (20.8) | 141 (79.2) | Reference | Reference |
|  | Yes | 16 (43.2) | 21 (56.8) | 0.34 (0.16 to 0.73)* | 0.30 (0.09 to 0.94)* |
| If I completely follow the Government's advice, I will lose touch with my friends and relatives | 5-point scale, 1=strongly disagree to 5=strongly agree | N=53, M=1.96, SD=1.16 | N=161, M=2.25, SD=1.26 | 1.23 (0.94 to 1.61) | 1.22 (0.83 to 1.78) |
| My friends or family will disapprove if I don't follow the Government's advice | 5-point scale, 1=strongly disagree to 5=strongly agree | N=52, M=3.92, SD=1.19 | N=159, M=4.05, SD=0.90 | 1.14 (0.83 to 1.56) | 1.17 (0.76 to 1.81) |
| If I don't follow the Government's advice, I could get in trouble with the police | 5-point scale, 1=strongly disagree to 5=strongly agree | N=53, M=3.98, SD=0.84 | N=159, M=3.83, SD=0.93 | 0.83 (0.58 to 1.18) | 0.88 (0.56 to 1.39) |
| If I follow the Government's advice, it will help save lives | 5-point scale, 1=strongly disagree to 5=strongly agree | N=54, M=4.54, SD=0.88 | N=161, M=4.39, SD=0.89 | 0.81 (0.55 to 1.19) | 0.71 (0.42 to 1.20) |
| If I follow the Government's advice, it will help protect the NHS | 5-point scale, 1=strongly disagree to 5=strongly agree | N=54, M=4.57, SD=0.66 | N=161, M=4.47, SD=0.81 | 0.82 (0.53 to 1.27) | 0.87 (0.50 to 1.53) |
| If I catch coronavirus, I may become very ill | 5-point scale, 1=strongly disagree to 5=strongly agree | N=53, M=4.51, SD=0.72 | N=156, M=4.45, SD=0.90 | 0.92 (0.63 to 1.34) | 1.02 (0.62 to 1.66) |
| If I catch coronavirus, it will have a severe impact on my family's wellbeing | 5-point scale, 1=strongly disagree to 5=strongly agree | N=52, M=4.15, SD=1.04 | N=158, M=4.18, SD=1.04 | 1.03 (0.76 to 1.39) | 1.34 (0.87 to 2.07) |
| If I leave home and meet other people, I could pass coronavirus to someone else | 5-point scale, 1=strongly disagree to 5=strongly agree | N=53, M=4.62, SD=0.56 | N=157, M=4.52, SD=0.75 | 0.79 (0.49 to 1.28) | 0.61 (0.29 to 1.25) |
| If I leave home and meet other people, I could catch coronavirus | 5-point scale, 1=strongly disagree to 5=strongly agree | N=54, M=4.74, SD=0.48 | N=161, M=4.5, SD=0.73 | 0.51 (0.29 to 0.92)* | 0.40 (0.16 to 0.97)* |
| If I follow the Government's advice it will have a negative impact on how much money I have | 5-point scale, 1=strongly disagree to 5=strongly agree | N=53, M=2.55, SD=1.29 | N=159, M=2.71, SD=1.28 | 1.11 (0.87 to 1.41) | 1.19 (0.87 to 1.64) |
| Because of the current lockdown there is more conflict between people that I live with | 5-point scale, 1=strongly disagree to 5=strongly agree | N=52, M=2.15, SD=1.13 | N=160, M=2.28, SD=1.27 | 1.08 (0.84 to 1.40) | 1.26 (0.86 to 1.86) |
| If I follow the Government's advice I will not be able to carry out important religious activities | 5-point scale, 1=strongly disagree to 5=strongly agree | N=50, M=2.58, SD=1.49 | N=148, M=2.69, SD=1.31 | 1.06 (0.84 to 1.35) | 1.08 (0.78 to 1.50) |
| I am enjoying spending more time at home during the lockdown | 5-point scale, 1=strongly disagree to 5=strongly agree | N=54, M=3.46, SD=1.21 | N=162, M=3.19, SD=1.22 | 0.82 (0.64 to 1.07) | 0.82 (0.58 to 1.17) |
| Because of coronavirus, I feel a sense of community with other people in my neighbourhood | 5-point scale, 1=strongly disagree to 5=strongly agree | N=54, M=2.98, SD=1.22 | N=162, M=3.25, SD=1.14 | 1.22 (0.94 to 1.60) | 1.50 (1.01 to 2.21)* |

\* $p \leq .05$
\*\* $p \leq .001$
† Adjusting for gender, age, having a child in the household, being extremely clinically vulnerable oneself, employment status, highest level of education or professional qualification, indices of multiple deprivation, social grade, living in a rural or urban area, living alone, marital status, and region.

#### Power

For analyses where symptoms were present in the household, we achieved 90% power to detect small effect sizes in logistic regression analyses (OR = 1.6,(16) α = .05, sample size n = 217, probability of having left the home = 0.75, one-tailed logistic regression). Using the same parameters, we achieved 83% power to detect small effect sizes using a two-tailed logistic regression. This is above the 80% threshold for statistical power considered acceptable.(17)

### No symptoms in household

Of those who reported that no-one in their household was symptomatic, 24.5% reported having gone out to shop for items other than groceries, toiletries or medicines (n = 476, 95% CI [22.6 to 26.4]), 5.9% reported meeting up with friends and/or family that they did not live with (n = 114, 95% CI [4.8 to 6.9]), and 4.3% reported having had visitors to their home in the last seven days (n = 81, 95% CI [3.4 to 5.3]). The mean number of outings made by participants was 6.77 (SD = 5.07, median = 6, mode = 0).

Personal and clinical factors (gender, age, having a child in the household, being extremely clinically vulnerable oneself, employment status, highest level of education or professional qualification, indices of multiple deprivation, social grade, living in a rural or urban area, living alone, marital status, and region [results for region not reported]) explained 8.0% of the variance in number of outings in the past week (see Table 4). More outings were made by males, those who reported working and those who lived in rural areas. Fewer outings were made by those who were clinically extremely vulnerable and those living in more deprived areas. Having a pet in addition to the above personal and clinical factors explained 9.1% of the variance in number of outings in the past week, with those who had a pet reporting making more outings in the past week.

**Table 4.**
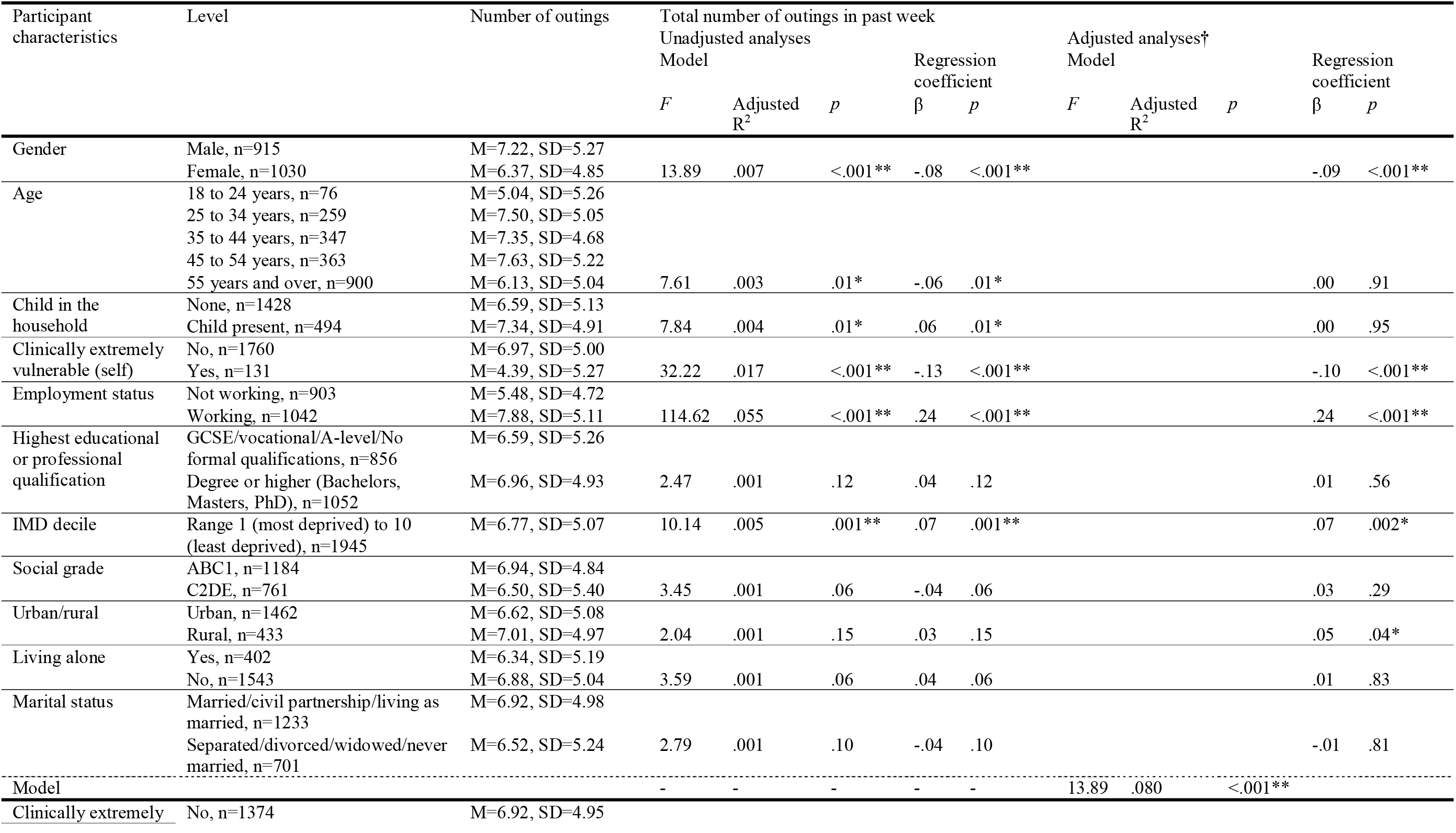
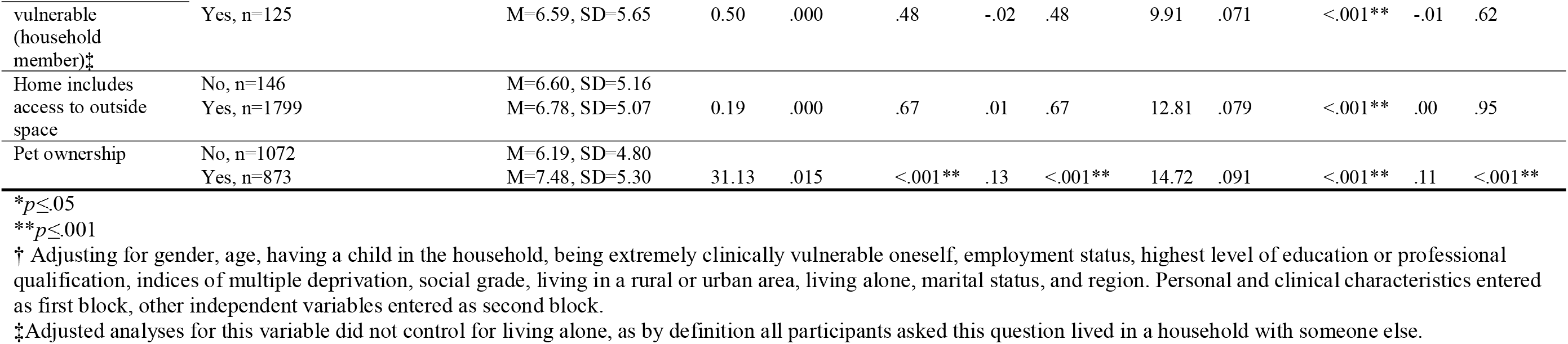
Associations between personal and clinical characteristics and total number of outings in past week in participants who reported no symptoms in the household.

Psychological and situational factors associated with more outings in the past week (Table 5) were: helping someone outside your household; decreased perceived effectiveness of Government measures to save lives and help protect the NHS; thinking that you would lose touch with friends and relatives if you followed Government advice; not enjoying spending more time at home during the lockdown; better self-reported general health; decreased perceived severity of COVID-19; decreased perceived likelihood of spreading COVID-19; decreased perceived legal consequences if not following Government advice; decreased perceived social pressure from friends and family to follow Government measures; knowing what Government measures were if no-one in the household was symptomatic; believing that you have had or currently have COVID-19; increased perceived financial cost of following Government measures; and decreased perceived social norms. Psychological and situational factors associated with fewer number of outings were receiving help from someone outside your household; decreased perceived impact of lockdown on physical health; reporting that you were self-isolating; increased worry about COVID-19; and increased perceived likelihood of catching COVID-19.

**Table 5.**
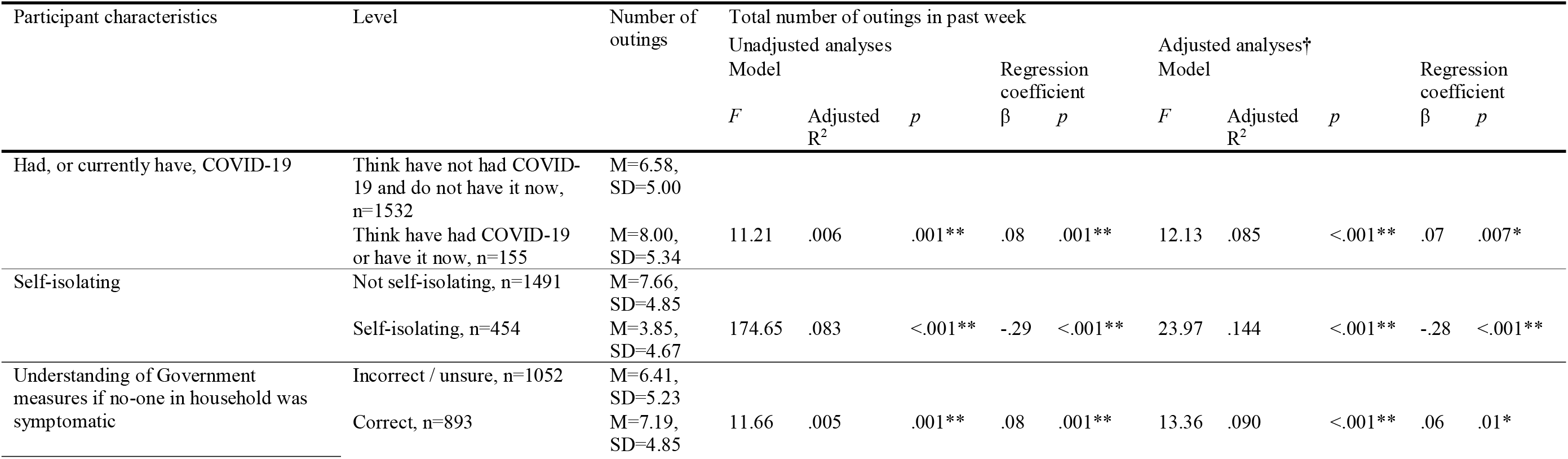
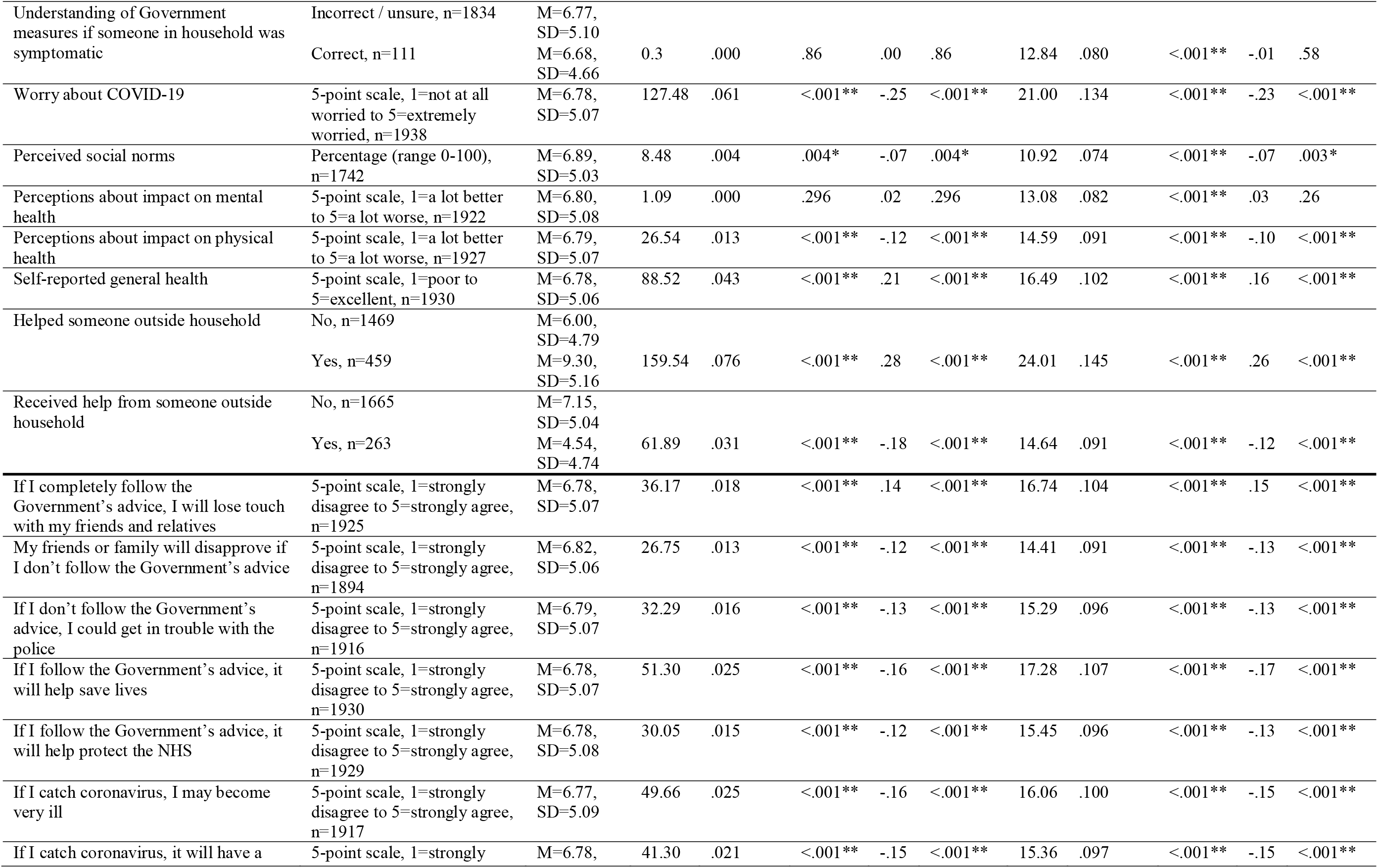
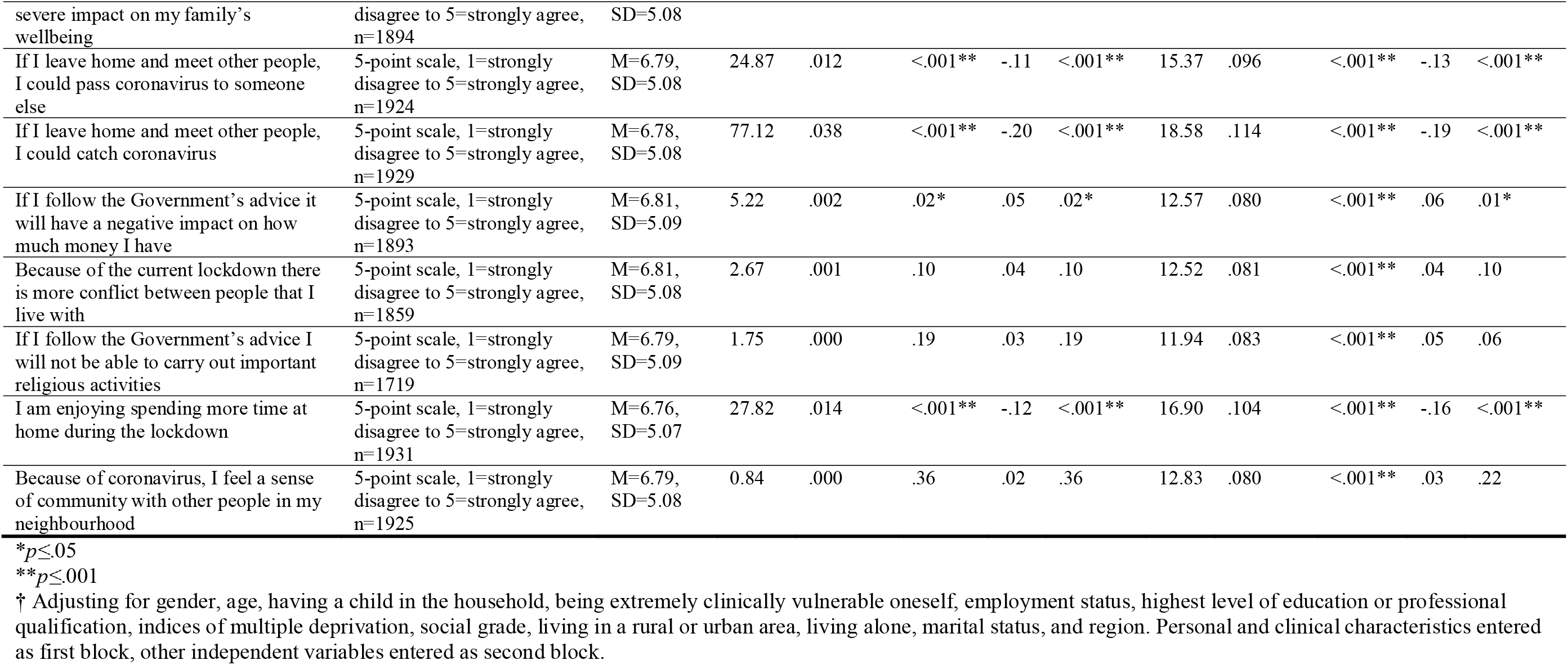
Associations between psychological and situational factors and total number of outings in past week in participants who reported no symptoms in the household.

Going out shopping for items other than groceries, toiletries or medicines in the past week was associated with being male, working and lower social grade (see Table 6).

**Table 6.**
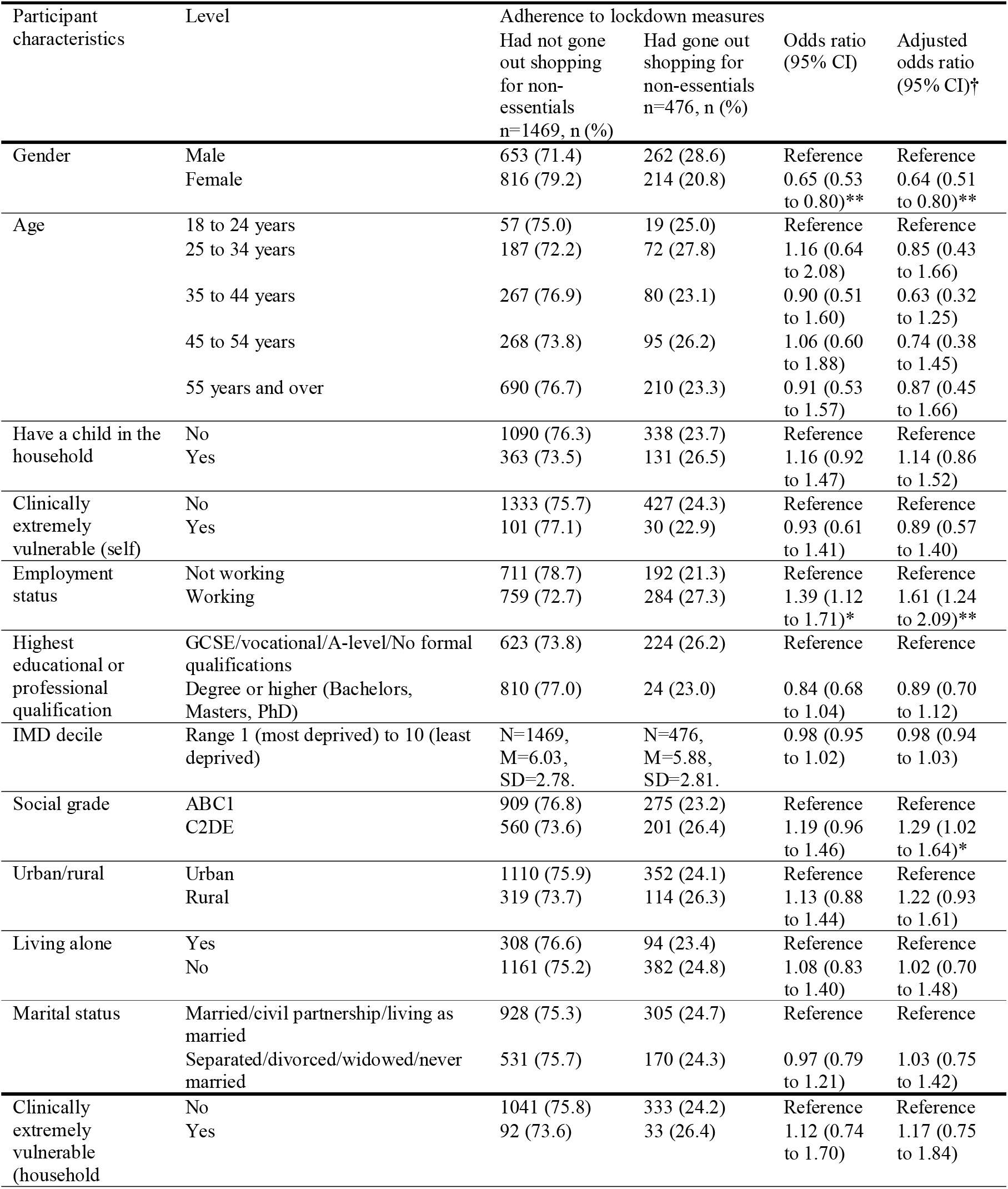
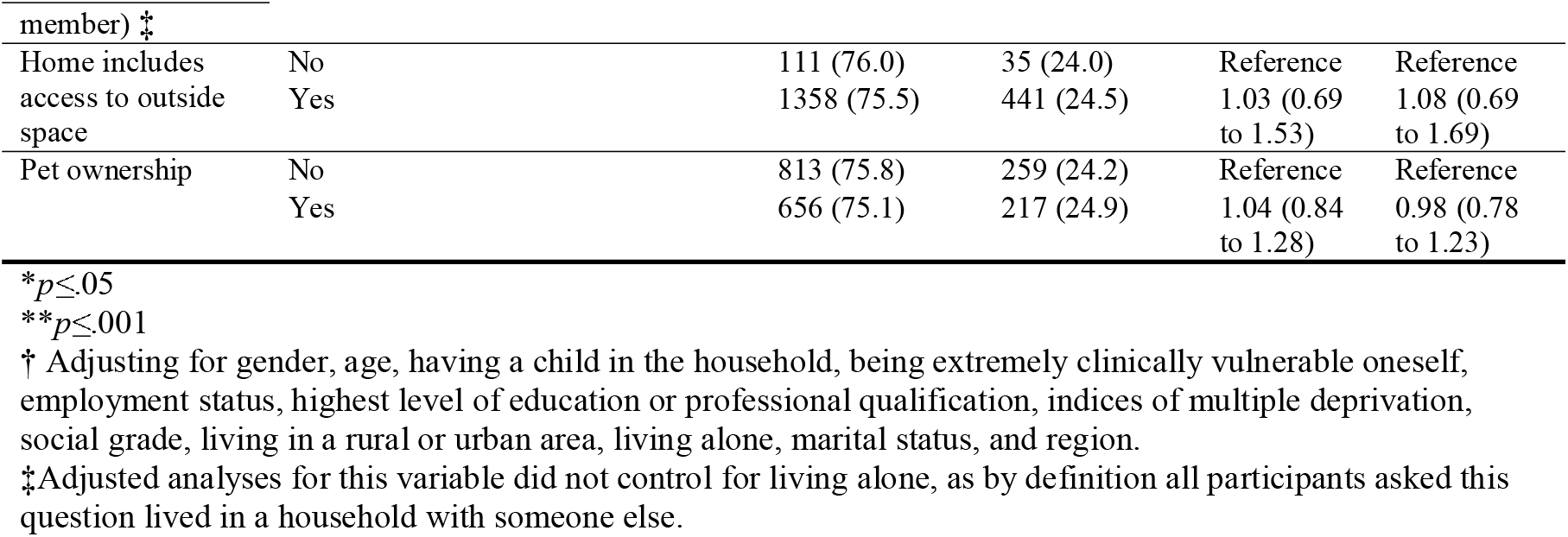
Associations between personal and clinical characteristics of participants who reported no symptoms in their household in the last week and having gone shopping for items other than groceries, toiletries or medicines (non-essentials).

Going out shopping for items other than groceries, toiletries or medicines in the past week was most strongly associated with thinking you have already had COVID-19 or have it now. Also associated with going out shopping for non-essentials were helping someone outside your household, thinking that you will lose touch with your friends or relatives if you follow Government guidance, and thinking that following Government guidance will negatively impact you financially (see Table 7). Not going out shopping for non-essentials in the past week was most strongly associated with having received help from someone outside your household in the last seven days. Also associated with not going out shopping for non-essentials were reporting that you were self-isolating, increased perceived likelihood of catching and spreading COVID-19, increased worry about COVID-19, increased perceived effectiveness of Government advice (saving lives and protecting the NHS), increased perceived severity of COVID-19 (to oneself and impact on family’s wellbeing), increased perceived disapproval from friends or family if you do not follow Government advice, increased perceived legal consequences of not following Government advice (e.g. getting into trouble with police), not knowing or being unsure about Government measures, and decreased perceived social norms.

**Table 7.** Associations between psychological and situational factors and having gone shopping for items other than groceries, toiletries or medicines (non-essentials) in the past seven days in participants who reported no symptoms in the household.

| Participant characteristics | Level | Adherence to lockdown measures |  | Odds ratio (95% CI) | Adjusted odds ratio (95% CI)† |
| --- | --- | --- | --- | --- | --- |
|  |  | Had not gone out shopping for non-essentials n=1469, n (%) | Had gone out shopping for non-essentials n=476, n (%) |  |  |
| Had COVID-19 | Think have not had COVID-19 | 1176 (76.8) | 356 (23.2) | Reference | Reference |
|  | Think have had COVID-19 | 104 (67.1) | 51 (32.9) | 1.62 (1.14 to 2.31)* | 1.73 (1.17 to 2.54)* |
| Self-isolating | Not self-isolating | 1098 (73.6) | 393 (26.4) | Reference | Reference |
|  | Self-isolating | 371 (81.7) | 83 (18.3) | 0.63 (0.48 to 0.81)** | 0.61 (0.45 to 0.84)* |
| Understanding of Government measures if no-one in household was symptomatic | Incorrect / unsure | 768 (73.0) | 284 (27.0) | Reference | Reference |
|  | Correct | 701 (78.5) | 192 (21.5) | 0.74 (0.60 to 0.91)* | 0.77 (0.61 to 0.97)* |
| Understanding of Government measures if someone in household was symptomatic | Incorrect / unsure | 1392 (75.9) | 442 (24.1) | Reference | Reference |
|  | Correct | 77 (69.4) | 34 (30.6) | 1.39 (0.92 to 2.11) | 1.27 (0.81 to 1.99) |
| Worry about COVID-19 | 5-point scale, 1=not at all worried to 5=extremely worried | N=1465, M=3.40, SD=0.97 | N=473, M=3.01, SD=1.00 | 0.67 (0.60 to 0.74)** | 0.66 (0.59 to 0.75)** |
| Perceived social norms | Percentage (range 0-100) | N=1312, M=74.19, SD=15.62 | N=431, M=70.35, SD=17.54 | 0.99 (0.98 to 0.99)** | 0.99 (0.98 to 0.99)** |
| Perceptions about impact on mental health | 5-point scale, 1=a lot better to 5=a lot worse | N=1452, M=3.43, SD=0.87 | N=470, M=3.43, SD=0.92 | 1.00 (0.89 to 1.13) | 1.01 (0.89 to 1.14) |
| Perceptions about impact on physical health | 5-point scale, 1=a lot better to 5=a lot worse | N=1457, M=3.15, SD=0.91 | N=470, M=3.11, SD=0.98 | 0.95 (0.85 to 1.07) | 0.95 (0.84 to 1.07) |
| Self-reported general health | 5-point scale, 1=poor to 5=excellent | N=1457, M=3.05, SD=1.06 | N=473, M=3.06, SD=1.01 | 1.01 (0.92 to 1.12) | 1.04 (0.93 to 1.17) |
| Helped someone outside household | No | 1138 (77.5) | 331 (22.5) | Reference | Reference |
|  | Yes | 321 (69.9) | 138 (30.1) | 1.48 (1.17 to 1.87)** | 1.56 (1.21 to 2.00)** |
| Received help from someone outside household | No | 1234 (74.1) | 431 (25.9) | Reference | Reference |
|  | Yes | 225 (85.6) | 38 (14.4) | 0.48 (0.34 to 0.69)** | 0.53 (0.36 to 0.79)* |
| If I completely follow the Government's advice, I will lose touch with my friends and relatives | 5-point scale, 1=strongly disagree to 5=strongly agree | N=1456, M=1.94, SD=1.04 | N=469, M=2.25, SD=1.18 | 1.28 (1.17 to 1.40)** | 1.30 (1.17 to 1.43)** |
| My friends or family will disapprove if I don't follow the Government's advice | 5-point scale, 1=strongly disagree to 5=strongly agree | N=1428, M=4.11, SD=0.93 | N=466, M=3.8, SD=1.05 | 0.73 (0.66 to 0.81)** | 0.72 (0.65 to 0.81)** |
| If I don't follow the Government's advice, I could get in trouble with the police | 5-point scale, 1=strongly disagree to 5=strongly agree | N=1448, M=3.98, SD=0.85 | N=468, M=3.77, SD=0.97 | 0.77 (0.69 to 0.87)** | 0.77 (0.69 to 0.88)** |
| If I follow the Government's advice, it will help save lives | 5-point scale, 1=strongly disagree to 5=strongly agree | N=1458, M=4.54, SD=0.72 | N=472, M=4.26, SD=0.95 | 0.67 (0.59 to 0.75)** | 0.66 (0.58 to 0.75)** |
| If I follow the Government's advice, it will help protect the NHS | 5-point scale, 1=strongly disagree to 5=strongly agree | N=1458, M=4.56, SD=0.74 | N=471, M=4.32, SD=0.90 | 0.71 (0.62 to 0.80)** | 0.71 (0.62 to 0.81)** |
| If I catch coronavirus, I may become very ill | 5-point scale, 1=strongly disagree to 5=strongly agree | N=1448, M=4.43, SD=0.82 | N=469, M=4.18, SD=0.94 | 0.73 (0.65 to 0.82)** | 0.72 (0.63 to 0.82)** |
| If I catch coronavirus, it will have a severe impact on my family's wellbeing | 5-point scale, 1=strongly disagree to 5=strongly agree | N=1431, M=4.12, SD=1.01 | N=463, M=3.86, SD=1.10 | 0.80 (0.72 to 0.88)** | 0.79 (0.71 to 0.88)** |
| If I leave home and meet other people, I could pass coronavirus to someone else | 5-point scale, 1=strongly disagree to 5=strongly agree | N=1453, M=4.44, SD=0.81 | N=471, M=4.14, SD=0.97 | 0.69 (0.62 to 0.78)** | 0.66 (0.58 to 0.75)** |
| If I leave home and meet other people, I could catch coronavirus | 5-point scale, 1=strongly disagree to 5=strongly agree | N=1459, M=4.45, SD=0.74 | N=470, M=4.14, SD=0.88 | 0.64 (0.56 to 0.72)** | 0.59 (0.52 to 0.68)** |
| If I follow the Government's advice it will have a negative impact on how much money I have | 5-point scale, 1=strongly disagree to 5=strongly agree | N=1426, M=2.47, SD=1.20 | N=467, M=2.64, SD=1.24 | 1.12 (1.03 to 1.22)* | 1.13 (1.03 to 1.24)* |
| Because of the current lockdown there is more conflict between people that I live with | 5-point scale, 1=strongly disagree to 5=strongly agree | N=1406, M=2.08, SD=1.15 | N=453, M=2.23, SD=1.16 | 1.11 (1.01 to 1.22)* | 1.08 (0.97 to 1.19) |
| If I follow the Government's advice I will not be able to carry out important religious activities | 5-point scale, 1=strongly disagree to 5=strongly agree | N=1294, M=2.59, SD=1.35 | N=425, M=2.70, SD=1.33 | 1.06 (0.98 to 1.15) | 1.08 (0.99 to 1.18) |
| I am enjoying spending more time at home during the lockdown | 5-point scale, 1=strongly disagree to 5=strongly agree | N=1458, M=3.29, SD=1.20 | N=473, M=3.21, SD=1.21 | 0.95 (0.87 to 1.03) | 0.94 (0.86 to 1.03) |
| Because of coronavirus, I feel a sense of community with other people in my neighbourhood | 5-point scale, 1=strongly disagree to 5=strongly agree | N=1455, M=3.36, SD=1.07 | N=470, M=3.30, SD=1.06 | 0.94 (0.86 to 1.04) | 0.99 (0.89 to 1.11) |

\* $p \leq .05$
\* $p \leq .001$
† Adjusting for gender, age, having a child in the household, being extremely clinically vulnerable oneself, employment status, highest level of education or professional qualification, indices of multiple deprivation, social grade, living in a rural or urban area, living alone, marital status, and region.

#### Power

For analyses where no symptoms were present in the household, we achieved 100% power to detect small effect sizes in logistic regression analyses (OR = 1.6, (16) α = .05, sample size n = 1945, probability of having gone out shopping for items other than groceries, toiletries or medicines = 0.25, one-tailed and two-tailed logistic regression). We achieved 94% power to detect small effect sizes in linear regression analyses (f^2^= 0.02,(17) α = .05, sample size n = 1945, number of tested predictors = 39, total number of predictors = 39).

## DISCUSSION

To the best of our knowledge, this is the first comprehensive study to investigate factors associated with self-isolation and behaviour during lockdown in the UK. In this sample, almost 10% of participants reported that either they or a household member had symptoms of COVID-19 (a cough or high temperature / fever) in the last week. Prevalence estimates by the UK Office for National Statistics indicate that at the time of data collection, 0.27% of the community population had COVID-19.(18) Nonetheless, Government regulations require all those with symptoms, or with symptoms in the household, to self-isolate. Our results suggest that adherence to this is poor. Three quarters of those with symptoms in their households reported that they had left their home in the past 24 hours. We also found no difference in out-of-home behaviour between those who did or did not have symptoms in their households. The UK will shortly enter a new phase of the pandemic, in which extensive testing, contact tracing and isolation will be required to keep the spread of COVID-19 in check (19). For this to succeed, adherence must be improved.

Our findings highlight several risk factors for poor adherence which could be targeted by public health messaging. Notably males were more likely to report symptoms. They were also more likely to report having been out in the last 24 hours if they or someone in their household was symptomatic, having gone out more times in the last week and shopping for items other than groceries, toiletries or medicines. Lower adherence among men was also noted in the UK during the 2009/10 H1N1 influenza pandemic.(10) Communication campaigns that specifically target men may therefore have merit.

Adherence with self-isolation was associated with increased worry about COVID-19 and increased perceived likelihood of catching COVID-19. As incidence declines, it is possible that worry will also decline, reducing adherence further. While it may be tempting to use fear-based messaging to combat this, this may influence other behaviours that the Government may wish to encourage, such as return to work.(20)

Adherence was also associated with having received help from someone outside your household in the last seven days. This makes intuitive sense – being able to ask someone else to run errands should reduce the need for you to leave home. Much has been made recently of the remarkable altruism of 750,000 people who signed-up to volunteer for the National Health Service, and the lack of jobs for them to do.(21) Allowing those in self-isolation to submit requests to this army of willing helpers may be a pragmatic way to improve adherence. Community volunteer networks, including neighbourhood groups on social media and “mutual aid” organisations.(22)

The finding that many of those with symptoms in the household believed themselves to be self-isolating, while also leaving their home, suggests that communication around self-isolation also requires much greater clarity.

Adherence to lockdown measures among those not reporting symptoms in their household was better, but still not perfect, with 75% reporting not going out to shop for non-essential items. Adherence to meeting up with friends or family from outside the household and not having visitors to the home was higher (94% and 95% respectively). Decreased adherence was associated with being male and working. It is plausible that those who are working may be more likely to be out and about for work and go to a shop for items other than groceries, toiletries, or medicines. Those working may also be more financially able to shop for nonessential items. Adherence was associated with factors associated with uptake of health behaviours, such as higher threat appraisals and positive appraisals of the coping response. These findings are in line with research from other countries.(23–25) Increased perceived social norms were also associated with adherence to lockdown measures.(23, 26) Other factors identified by a recent rapid review of adherence to quarantine,(9) such as lower perceived social pressure to adhere to measures and decreased perceived legal consequences of not following measures were also associated with non-adherence to lockdown measures. While perceiving greater negative financial consequences of Government measures was associated with non-adherence to lockdown measures, there was no longer evidence for an association after correcting for multiple adjustments. This is different from research finding decreased intention to adhere to quarantine measures in Israel.(27) Knowledge of UK Government guidance where no-one in the household was symptomatic was also associated with non-adherence to lockdown measures.(9) These findings suggest that improvement in adherence to lockdown measures is likely to be achieved by emphasising these are actions that most people are taking, that are having a positive impact, and that others around you want you to do. They also suggest threatening legal consequences for not engaging in the activity is less likely to be a strong motivator.

This study has several limitations. First, despite using quota sampling so that personal characteristics of the sample broadly reflected those in the UK general population, we cannot be sure that survey respondents are representative of the general population.(28, 29) Research during disasters and public health crises often require a trade-off between rapidity and rigour.(11) The use of a professional market research agency using industry standard opinion polling methodology provides a suitable balance in these circumstances. Second, all data were self-reported and may have been susceptible to social desirability bias.(30) However, preliminary data indicate that self-reported physical distancing is associated with real-world behaviour.(31) If anything, the rates of adherence we observed may be over-estimates of adherence. Third, we did not ask participants if they came into close contact (within 2 metres) with anyone from another household while they were out and about. We cannot tell if those who went out more, including shopping for non-essentials, were in close contact with more people than those who went out less or who did not go out shopping for non-essentials. However, we reason that those who have been out and about more times have a greater chance of having come into close contact with someone from outside their household. This is only relevant for those who reported no symptoms in their household. For those who reported symptoms in their household, any outing is a breach of guidance. Fourth, we used a cumulative measure of ‘outings’ for our outcome measure. It is possible that participants who went to a supermarket for groceries or toiletries may also have bought other non-essential items, and reported this as one outing to buy groceries, toiletries, or medicines, and one outing to buy other items. Thus, in our calculations, a single trip could have counted as two outings. Our variable indicating the number of outings made in the past week, and the number of outings made for non-essential items specifically may therefore be over-estimates. Fifth, the cross-sectional nature of data collection means we are unable to draw causal inferences. Sixth, although the total sample size was large, a small percentage of the population reported that they or someone in their household had experienced symptoms of COVID-19 in the last week. Therefore, the sample size for analyses investigating associations with self-isolation was small, resulting in decreased power and wide confidence intervals. Despite all analyses achieving over 83% power, these results should be taken with caution.

To the best of our knowledge, this is the first study investigating factors associated with adherence to self-isolation and lockdown measures in the UK. Data suggest that self-reported adherence to self-isolation measures was poor. This has important implications for policies that attempt to prevent the spread of COVID-19 through self-isolation, such as contact tracing, and will become more prominent when restrictions on movement are eased further. Psychological factors including perceived effectiveness of lockdown measures, should be emphasised in communications. Effective use of volunteer programmes and help within the neighbourhood or community may also improve adherence to self-isolation and lockdown measures.

## Data Availability

Anonymised data will be made available upon reasonable request.

## FUNDING SOURCES

LS, RA and GJR are supported by the National Institute for Health Research Health Protection Research Unit (NIHR HPRU) in Emergency Preparedness and Response, a partnership between Public Health England, King’s College London and the University of East Anglia. RA, HL, IO and CR are supported by the NIHR HPRU in Behavioural Science and Evaluation, a partnership between Public Health England and the University of Bristol. CR is also supported by the NIHR HPRU in Emerging and Zoonotic Infections and NIHR HPRU in Gastrointestinal Infections. The views expressed are those of the authors and not necessarily those of the UKRI, NIHR, Public Health England or the Department of Health and Social Care. The NIHR HPRU Emergency Preparedness and Response funded the study. An MRC award under the MRC COVID-19 Rapid Response call (grant number MC_PC_19071) funded RA, HL, IO, CR, LY and GJR’s time.

## TRANSPARENCY DECLARATION

The authors affirm that the manuscript is an honest, accurate, and transparent account of the study being reported; that no important aspects of the study have been omitted; and that any discrepancies from the study as originally planned have been explained.

## DATA SHARING STATEMENT

Anonymised data will be made available upon reasonable request.

## AUTHOR CONTRIBUTION STATEMENT

The study was conceptualised by RA, HL, IO, CR, LY and GJR. LS completed all analyses, using data from YouGov Plc. All authors contributed to, and approved, the final manuscript. For any enquiries about the data in this report please contact King’s College London.

## LICENSE

The Corresponding Author has the right to grant on behalf of all authors and does grant on behalf of all authors, a worldwide licence (the BMJ license) to the Publishers and its licensees in perpetuity, in all forms, formats and media (whether known now or created in the future), to i) publish, reproduce, distribute, display and store the Contribution, ii) translate the Contribution into other languages, create adaptations, reprints, include within collections and create summaries, extracts and/or, abstracts of the Contribution and convert or allow conversion into any format including without limitation audio, iii) create any other derivative work(s) based in whole or part on the on the Contribution, iv) to exploit all subsidiary rights to exploit all subsidiary rights that currently exist or as may exist in the future in the Contribution, v) the inclusion of electronic links from the Contribution to third party material where-ever it may be located; and, vi) licence any third party to do any or all of the above. All research articles will be made available on an open access basis (with authors being asked to pay an open access fee—see <u>copyright, open access, and permission to reuse</u>). The terms of such open access shall be governed by a <u>Creative Commons</u> licence—details as to which Creative Commons licence will apply to the research article are set out in our worldwide licence referred to above.

## DISSEMINATION DECLARATION

Dissemination of survey results to participants is not possible due to the anonymous nature of data collection.

## REFERENCES

1. Cabinet Office. New rules on staying at home and away from others. 23 March 2020.

2. Johnson B. PM address to the nation on coronavirus: 23 March 2020. Available from: https://www.gov.uk/government/speeches/pm-address-to-the-nation-on-coronavirus-23-march-2020.

3. NHS [internet]. Check if you have coronavirus symptoms 2020 [updated 18 May 2020; accessed 25 May 2020]. Available from: https://www.nhs.uk/conditions/coronaviruscovid-19/check-if-you-have-coronavirus-symptoms/.

4. Dominic Raab. Foreign Secretary’s statement on coronavirus (COVID-19). 16 April 2020. Available from: https://www.gov.uk/government/speeches/foreign-secretarysstatement-on-coronavirus-covid-19-16-april-2020.

5. Johnson B. PM statement in the House of Commons. 11 May 2020. Available from: https://www.gov.uk/government/speeches/pm-statement-in-the-house-of-commons-11-may-2020.

6. HM Government. Our plan to rebuild: The UK Government’s COVID-19 recovery strategy. 2020.

7. Smith LE, Mottershaw AL, Egan M, Waller J, Marteau TM, Rubin GJ. The impact of believing you have had COVID-19 on behaviour: Cross-sectional survey. medRxiv. 2020:2020.04.30.20086223.

8. Rogers RW, Prentice-Dunn S. Protection motivation theory. 1997.

9. Webster RK, Brooks SK, Smith LE, Woodland L, Wessely S, Rubin GJ. How to improve adherence with quarantine: rapid review of the evidence. Public Health. 2020;182:163–9.

10. Rubin GJ, Amlot R, Page L, Wessely S. Public perceptions, anxiety, and behaviour change in relation to the swine flu outbreak: cross sectional telephone survey. BMJ. 2009;339:b2651.

11. Rubin GJ, Amlot R, Page L, Wessely S. Methodological challenges in assessing general population reactions in the immediate aftermath of a terrorist attack. Int J Methods Psychiatr Res. 2008;17 Suppl 2:S29–35.

12. Rubin GJ, Brewin CR, Greenberg N, Simpson J, Wessely S. Psychological and behavioural reactions to the bombings in London on 7 July 2005: cross sectional survey of a representative sample of Londoners. BMJ. 2005;331(7517):606.

13. Rubin GJ, Page L, Morgan O, Pinder RJ, Riley P, Hatch S, et al. Public information needs after the poisoning of Alexander Litvinenko with polonium-210 in London: cross sectional telephone survey and qualitative analysis. BMJ. 2007;335(7630):1143.

14. Ware JE, Jr., Sherbourne CD. The MOS 36-item short-form health survey (SF-36). I. Conceptual framework and item selection. Med Care. 1992;30(6):473–83.

15. Rubin GJ, Smith LE, Melendez-Torres GJ, Yardley L. Improving adherence to ‘Test, Trace and Isolate’. BMJ. Submitted.

16. Chen HN, Cohen P, Chen S. How Big is a Big Odds Ratio? Interpreting the Magnitudes of Odds Ratios in Epidemiological Studies. Commun Stat-Simul C. 2010;39(4):860–4.

17. Cohen J. Statistical power analysis for the behavioral sciences. 2nd ed. Hillsdale, N.J.; Hove: Erlbaum Associates; 1988. xxi, 567 p. p.

18. Office for National Statistics. Coronavirus (COVID-19) Infection Survey pilot: England. 14 May 2020 [updated 14 May 2020; accessed 25 May 2020]. Available from: https://www.ons.gov.uk/peoplepopulationandcommunity/healthandsocialcare/conditionsanddiseases/bulletins/coronaviruscovid19infectionsurveypilot/england14may2020.

19. Hellewell J, Abbott S, Gimma A, Bosse NI, Jarvis CI, Russell TW, et al. Feasibility of controlling COVID-19 outbreaks by isolation of cases and contacts. Lancet Glob Health. 2020;8(4):e488–e96.

20. Rubin GJ, Potts HWW, Michie S. The impact of communications about swine flu (influenza A H1N1v) on public responses to the outbreak: results from 36 national telephone surveys in the UK. Health Technol Asses. 2010;14(34):183–266.

21. Butler P. NHS coronavirus crisis volunteers frustrated at lack of tasks. The Guardian. 2020 3 May 2020.

22. Booth R. Community aid groups set up across UK amid coronavirus crisis. The Guardian. 2020 16 March 2020.

23. Gouin J-P, MacNeil S, Switzer A, Carrese-Chacra E, Durif F, Knauper B. Social, Cognitive, and Emotional Predictors of Adherence to Physical Distancing During the COVID-19 Pandemic. PsyArXiv. 2020.

24. Pollack Y, Dayan H, Shoham R, Berger I. Predictors of adherence to public health instructions during the COVID-19 pandemic MedRxiv. 2020.

25. Seale H, Heywood AE, Leask J, Sheel M, Thomas S, Durrheim DN, et al. COVID-19 is rapidly changing: Examining public perceptions and behaviors in response to this evolving pandemic. MedRxiv. 2020.

26. Goldberg MH, Gustafson A, Maibach EW, van der Linden S, Ballew MT, Bergquist P, et al. Social norms motivate COVID-19 preventive behaviours. PsyArXiv. 2020.

27. Bodas M, Peleg K. Self-Isolation Compliance In The COVID-19 Era Influenced By Compensation: Findings From A Recent Survey In Israel. Health Aff (Millwood). 2020:101377hlthaff202000382.

28. Office for National Statistics. Internet users, UK: 2019. 2019.

29. Wright KB. Researching Internet-Based Populations Advantages and Disadvantages of Online Survey Research, Online Questionnaire Authoring Software Packages, and Web Survey Services. J Comput-Mediatd Commun. 2005; 10(3). https://onlinelibrary.wiley.com/doi/full/10.1111/j.1083-6101.2005.tb00259.x.

30. Daoust J-F, Nadeau R, Dassonneville R, Lachapelle E, Belanger E, Savoie J, et al. How to survey citizens’ compliance with COVID-19 public health measures? Evidence from three survey experiments. SocArXiv. 2020.

31. Gollwitzer A, Martel C, Marshall J, Hohs JM, Bargh JA. Connecting self-reported social distancing to real-world behavior at the individual and U.S. State level. PsyArXiv. 2020.

